# Altered DNA methylation profiles in blood from patients with sporadic Creutzfeldt-Jakob disease

**DOI:** 10.1101/2020.08.25.20181495

**Authors:** Luke Dabin, Fernando Guntoro, Tracy Campbell, Tony Bélicard, Adam R. Smith, Rebecca G. Smith, Rachel Raybould, Jonathan M. Schott, Katie Lunnon, Peter Sarkies, John Collinge, Simon Mead, Emmanuelle Viré

## Abstract

Prion diseases are fatal and transmissible neurodegenerative disorders caused by the misfolding and aggregation of prion protein. Although recent studies have implicated epigenetic variation in common neurodegenerative disorders, no study has yet explored their role in human prion diseases. Here we profiled genome-wide blood DNA methylation in the most common human prion disease, sporadic Creutzfeldt-Jakob disease (sCJD). Our case-control study (n=219), when accounting for differences in cell type composition between individuals, identified 38 probes at genome-wide significance (p < 1.24×0^-7^). Nine of these sites were taken forward in a replication study, performed in an independent case-control (n=186) cohort using pyrosequencing. Sites in or close to *FKBP5, AIM2* (2 probes), *UHRF1, KCNAB2, PRNP, ANK1* successfully replicated. The blood-based DNA methylation signal was tissue- and disease-specific, in that the replicated probe signals were unchanged in case-control studies using sCJD frontal-cortex (n=84), blood samples from patients with Alzheimer’s disease, and from inherited and acquired prion diseases. Machine learning algorithms using blood DNA methylation array profiles accurately distinguished sCJD patients and controls. Finally, we identified sites whose methylation levels associated with prolonged survival in sCJD patients. Altogether, this study has identified a peripheral DNA methylation signature of sCJD with a variety of potential biomarker applications.

## Introduction

Human prion diseases are typically rapidly progressive neurodegenerative conditions associated with misfolding of prion protein (PrP)[10]. They include sporadic, Mendelian genetic, and acquired disorders which present and progress heterogeneously, although all are inevitably fatal. Neuropathologically, the diseases are characterized by spongiform changes in the gray matter with neuronal loss, reactive gliosis, and the accumulation of misfolded forms of PrP. The causative and transmissible agent of prion diseases, or prion, is thought to comprise solely or predominantly of misfolded forms or PrP forming paired double helical fibrils[56]. Mechanisms inspired by the prion concept are now widely adopted in neurodegenerative diseases associated with misfolded forms of other proteins and peptides[10].

The most common form of human prion disease is sporadic CJD (sCJD), occurring almost exclusively in adults over the age of 40 with an annual incidence of ~2 per million population. Whilst sCJD occurs seemingly at random in the population, most of the cases present between 60 and 80 years old. Prion diseases are under strong genetic control, with the most powerful risk factors being located at the PrP gene locus *(PRNP)*, particularly the polymorphism at codon 129 in the *PRNP* gene where methionine (~60% allele frequency in Europeans) or valine is encoded and both homozygous genotypes are at increased risk of disease[45,11]. Codon 129 genotype also modifies the incubation period of acquired prion diseases and the resulting clinico-pathological phenotype[12,49]. Disease severity, as measured by speed of decline in cognitive function, is fastest in those with the methionine homozygous genotype, and slowest in those with the heterozygous genotype [47,35].

According to epidemiological case definitions, a brain biopsy or post-mortem examination is necessary to confirm a diagnosis of definite sCJD, although neurological investigations such as cerebrospinal fluid (CSF) analysis, Magnetic Resonance Imaging (MRI), Electroencephalogram (EEG) and *PRNP* analysis can lead to a confident pre-mortem diagnosis of sCJD once it is suspected by specialist physicians. More recently, extensive research has been directed towards identification of specific and selective biomarkers such as metabolites or proteins. To date, these efforts have largely concentrated on altered protein concentrations, or the real-time quaking induced conversion assay using CSF[59]. The need for biomarkers in easily accessible tissues such as blood is important because such tests could help make diagnoses and prognoses earlier and screen individuals before invasive procedures, tissue or blood donation. Such approaches might also improve screening of individuals for inclusions in clinical trials prior to irretrievable neuronal damage.

Epigenetic signals are emerging as biomarkers for screening and early detection of various diseases, for prognostic and treatment monitoring, and for predicting future risk of disease development[15]. Accumulating evidence suggests that epigenetic modification of gene expression regulates memory acquisition and consolidation in the healthy brain and that epigenetic dysregulation contributes to the impaired cognition and neuronal death that are associated with neurodegenerative diseases[23]. Whether these changes are causally involved in diseases remain poorly understood. Preliminary analyses identified common changes in DNA methylation in several neurodegenerative diseases [51] pointing to shared regulatory programs. However, the contribution of epigenetic mechanisms to the initial steps and disease progression in neurodegeneration is yet still poorly understood.

Here, we compared genome-wide DNA methylation profiles in whole blood taken from patients with sCJD and age-matched healthy controls and characterized the genomic distribution of differentially methylated sites and regions. We identified sites where loss of DNA methylation correlates with disease progression. We further demonstrated that the DNA methylation signature is not altered in AD or in other prion diseases and found that the sites affected in blood are not differentially methylated in brain. Next, we show that machine learning models trained using DNA methylation profiles can discriminate sCJD from controls individuals. Finally, our findings, when used in combination with the genetic information of the patients, help refine disease duration predictions. We report the first sCJD DNA methylation-based blood signature that provides diagnostic and genotype independent prognostic information.

## Methods

### Patient samples and genomic DNA extraction

Patients with definite diagnosis of sCJD according to World Health Organization criteria were recruited by the National Prion Clinic (London, UK), and other referrers in the United Kingdom between 1995 and 2018. All sCJD patients were of UK residency. Blood or DNA from control donors was sourced from Cardiff University (Cardiff, UK), or from the National Prion Clinic (London, UK). Genomic DNA was extracted from peripheral blood using either a BACC2 DNA extraction kit (GE Healthcare, IL, USA) or a Zymo Quick gDNA MiniPrep Kit (Zymo Research, CA, USA) according to the manufacturers’ instructions.

GPower 3.1 [15] was used to estimate sample sizes required to power a genome-wide study. As the magnitudes of differences in blood-derived DNA methylation between sCJD and controls were unknown, Cohen’s δ was used as a measure of differences between means. To detect a difference informally described by Cohen as “small” (δ = 0.3) with a 95% power and a 0.05 error rate, 111 sCJD cases and 111 healthy controls were needed[9]. For pyrosequencing assays, post-hoc power calculations revealed that replication of effects observed in the discovery phase required between 7 and 50 sCJD and control samples to reach 95% power. Where brain samples were used, genomic DNA was extracted from frontal cortex grey matter from 51 autopsied sCJD brains in Biosafety Level 3 facilities. 50-100 mg of tissue was transferred to a 2 ml screw-capped tube (Eppendorf, Germany) and incubated in 450 μl ATL lysis buffer (Qiagen, NL) and 50 μl proteinase K (from 20 mgml^-1^ stock) in a Thermomixer Comfort heating block (Eppendorf, Germany) overnight at 50°C with mixing at 800 rpm. The next day samples were mixed with 500 μl of TRIS equilibrated phenol (Sigma-Aldrich, DE) by inversion. Tubes were then centrifuged at 16,100 *g* for 5 mins at room temperature before the upper aqueous phase was transferred to a fresh tube and lower organic phase was discarded into a phenol waste bottle. Addition of TRIS-equilibrated phenol, centrifugation and selection of the aqueous phase was repeated before 500 μl of a 1:1 v/v TRIS-equilibrated phenol and chloroform mixture was added and mixed by inversion. Centrifugation and selection of the aqueous phase was repeated before a final addition of 500 μl chloroform to the sample, which was centrifuged at 16,100 *g* for 2 mins. The upper aqueous phase was transferred to a clean tube and from containment level 3 facilities to containment level 2 facilities, where 500 μl 100% ethanol was added and mixed to induce DNA precipitation. DNA was spooled out onto a flame-sealed glass Pasteur pipette and left to dry for 2 mins, before resuspension in a 1.5 ml Eppendorf tube containing 500 μl Tris-EDTA buffer (Sigma-Aldrich, DE). Genomic DNA from was similarly extracted from 33 non-prion control frontal cortex samples acquired from Cambridge Brain Bank (University of Cambridge, UK). Concentration of extracted DNA was measured via Qubit (Thermofisher, MA, USA) and integrity was assessed using gDNA Tapestation ScreenTapes (Agilent, CA, USA). Samples with a DIN<7.0 were excluded from the study. Ethical approval was obtained from the National Hospital Local Research Ethics Committee.

### Genome-wide DNA methylation profiling

Bisulphite conversion of 500ng of genomic DNA was performed using the Zymo EZ-96 DNA Methylation-Gold Kit™ (Zymo, CA, USA) according to the manufacturer’s instructions. All DNAs were hybridised onto the Infinium ® Human Methylation 450K BeadChip array (Illumina, CA, USA). Fully methylated and unmethylated DNA standards (Zymo, CA, USA) and a commercially available leukocyte-derived DNA standard (AMSBIO, UK) were included as comparative controls for extreme variance of global DNA methylation profiles, while control probes on the array were used to monitor bisulphite conversion efficiency.

### Identification of differentially methylated CpG loci

Analyses were performed in RStudio version 1.0.136 using R v3.4.1. IDAT files were loaded into ChAMP version 2.10.2 [39] and normalised using the BMIQ method[57]. Reported values at thousands of 450K array probes are known to associate with which position on the beadchip the sample was assigned[25]. We confirmed this using Singular Value Composition and used ComBat[26] to correct for Beadchip number and sample position batch effects. Leukocyte population heterogeneity was estimated and corrected for using the Houseman method[22]. Quantile-quantile plots and Manhattan plots were generated using an in-house script adapted from qqman version 0.1.4 [60] and the pQQ function from version 7.0.0 of the haplin package[19]. An area of 95% confidence level was shaded around the reference line in the QQ plot. For Manhattan plots, a significance threshold was drawn at Bonferroni-adjusted significance threshold of p-value =1.24×10^-7^. Principal component analysis (PCA) was performed using the R packages FactoMineR version 1.41 [30] and factoextra version 1.0.5 (https://CRAN.R-proiect.org/package=factoextra). Heatmaps were produced using a script adapted from (https://github.com/obigriffith/) using either the top 38 or 1000 most significant differentially methylated CpG loci. Hierarchical clustering analysis was performed using the average clustering method based on Euclidean distance. Plotly (https://plot.ly/) was used to generate the pie charts showing the genomic locations of the CpG loci.

We used limma version 3.36.5 to build linear regression models of methylation versus disease status with age and sex included as covariates[48]. Bivariate Pearson’s correlations were used to correlate identified DMPs with MRC Scale score and slope in sCJD patients, while a one-way ANOVA with Dunnett’s post-hoc test was used to test for association of methylation at DMPs with genotype at codon 129 of *PRNP*. To identify DMRs, we used the DMRcate function in ChAMP [24] setting the lambda at 500. We used Bioconductor package ‘PWMEnrich’ [54] to perform motif scanning and enrichment analysis on the DMPs using probe sequences extracted from the Illumina HumanMethylation450K manifest file, and on the DMRs using sequences retrieved from Bioconductor package BSgenome.Hsapiens.UCSC.hg19 version 1.4.0[55]. For pathways and ontology analysis MetaCore[37] was used. Differentially methylated probes (bonferroni corrected *P* ≤ 0.1) and their Δβ values were uploaded to MetaCore and analysed as a single experiment, using the Illumina 450K array background and a significance threshold of *P* ≤ 0.1. Pathmap analysis was performed using Dijkstra’s shortest path algorithm with a maximum node distance of 2, using canonical pathways

### Pyrosequencing

Genomic DNA was bisulphite converted as described above. PCR primers were designed using the PyroMark® Assay Design 2.0 software (Qiagen, NL) and manually adjusted using the following criteria: (i) Amplicon length less than 200 bp, (ii) Sequencing read length below 40 nt, (iii) Forward and reverse primers have a length of 20-25 nt and do not overlap CpG sites, (iv) Sequencing primer does not exceed 20 nt in length and has an optimal T^m^ of 40°C. Primers were checked for specificity *in silico* using BiSearch [2] and optimum annealing temperatures were determined using PCR with a annealing temperature gradient between 52-62°C. PCR amplification of bisulphite-converted DNA was performed in a Veriti 96-Well Thermo Cycler (Thermofisher, MA, USA) using the following mastermix per 1 μl bisulphite-converted DNA: 1X Buffer B1 (Solis BioDyne, EE), 25 mM MgCl2 (Solis BioDyne, EE), 10 mM dNTP mix (Promega, WI, USA), 10 μM forward/reverse primer mix (Supplementary Table 5, online resource), 1 U HotFire Polymase (Solis BioDyne, EE) and RNAse-free water (Thermofisher, MA, USA) to a total reaction volume of 20 μl. PCR products were sequenced using the Pyromark™ Q96 system (Qiagen, NL) according to the manufacturer’s protocol. Where the assay permitted, a non-CpG cytosine was selected as a control for complete bisulphite conversion. Statistical analysis was performed in SPSS version 25 using a linear regression model of methylation (%) versus disease status, with age and sex as covariates. Pyrosequencing data are presented as Tukey boxplots, where the box is divided by the median and extend across the interquartile range. Whiskers protrude from the box up to 1.5 times the interquartile range, and outlying values are plotted as single points behind it.

### Cortisol profiling

500 μl aliquots of sera taken from patients and controls between 10am and 4pm were sent to The Doctor’s Laboratory (Sonic Healthcare Ltd., London, UK), where cortisol concentration was measured using the Elecsys Cortisol II assay (Roche: 11875116160) on a Cobase 801 module. Difference in cortisol concentration between sCJD and control donors was calculated in SPSS version 25 using a linear regression model with age and sex as covariates.

### Machine learning classification

Preprocessing of the β values from the 1000 most significantly altered loci from 105 sCJD and 105 control patients was performed in R version 3.5.1 and Rstudio version 1.1.456. The deep learning neural network model was created using the machine learning Tensorflow version 1.12.0 library [34](Abadi et al., 2015) run in Python 3 platform. The dataset was randomised and partitioned into training and test sets in a 50:50 ratio. The architecture of the model consisted of three dense hidden layers using the rectified linear unit activation function, and a sigmoid output layer to compute a score between 0 and 1 for classifying sCJD status. Furthermore, L2 weight regularization of 1×10^-3^ was applied to the hidden layers to impose a penalty to the cost function and reduce overfitting. The model was compiled using the Adam optimizer [13] with a learning rate of 5×10^-5^. Classification model performance was measured using two different metrics; accuracy and binary cross-entropy loss. The model was then fit into the training and test sets in 400 epochs with a batch size of 8. The model was compared to a basic random forest classifier imported from version 0.21 of the Python ‘sklearn’ library[46], which was also used to analyse the ROC curve, AUC and 10-k fold validation.

### Kaplan Meier curves

Survival curves were generated using the packages ggplot2 and survminer [28] using the ggsurvplot() function. Survival curves were plotted based on data from 102 sCJD patients after removing 12 patients without complete clinical information. For each of the 38 DMPs, beta values from each patient were categorized into “High” or “Low” according to the mean methylation levels. The confidence interval and p-values were computed using default options (log-rank test for p-value). For the Codon129 MM-MV-VV comparisons a threshold p-value<0.05 was used so that there is a significant difference within the particular subgroup i.e. methylation level has an effect. For the effect of DNA methylation on survival independent of codon 129, “High versus Low”, p-value>0.05 was used. For the combined survival analysis, beta values from the DMPs were transformed into z-scores after passing parametric test (Shapiro-Wilk cg01084918 p-value=0.4967; cg05343106 p-value=0.4372; cg17641710 p-value=0.2549). The average z-score is then used to inform levels of methylation (z-score >0, “High”; z-score <0, “Low”).

## Results

### DNA methylation profiles are altered in blood from sCJD patients

We profiled DNA methylation in whole blood from UK sCJD (n=114) patients and age and sex-matched controls (n = 105; Table 1) using Illumina Infinium HumanMethylation450 BeadChip arrays (450K arrays). One sample and 82,158 probes were discarded, which either failed quality control or were included in published blacklists [43,39] (Supplementary Fig. 1a, online resource). To identify an association between methylation β values[4] and sCJD disease status, we used a mixed linear regression model, including age and sex as covariates. Principal component analysis (PCA) of pre-processed data showed that the first two components were highly correlated with the contrasted groups, accounting for 9.7% and 6.7% of the variability respectively (Fig. 1a and Supplementary Fig. 1b, online resource), demonstrating that stringent quality control and adjustment steps removed sources of variation from factors unrelated to the biological variables. In this initial analysis, uncorrected for cell composition, we found 22,398 differentially methylated positions (DMPs) between healthy controls and sCJD patients (Bonferroni correction; p < 1.24×10^-7^) (Fig. 1b). Of those, 283 sites showed an absolute change in methylation greater than 10% (Δβ >|0.11) in sCJD (Fig. 1 b and Supplementary Table 1, online resource).

**Table 1.** Sample characteristics of the individuals included in the analyses.

| **Study Stage** | **Group** | **N** | **Average Age (range)** | **Sex (% F)** | **Codon 129 (%)**<br>**MM:MV:VV** | **Average MRC Scale Score (range)** |
| --- | --- | --- | --- | --- | --- | --- |
| Exploratory | sCJD | 114 | 67.6 (49-85) | 50.9 | 46:22:32 | 6.3 (0-20) |
|  | Control | 106 | 69.3 (41-83) | 55.7 | 44:43:13 | 20 |
| Replication | sCJD | 72 | 67.3 (26-86) | 58.3 | 54:23:23 | 4.5 (0-18) |
|  | Control | 114 | 78.2 (61-93) | 64.9 | Unknown | 20 |
| Specificity | Control | 114 | 78.2 (61-93) | 64.9 | Unknown | 20 |
|  | AD | 60 | 72.8 (70-77) | 56.7 | Unknown | Unknown |
|  | iCJD | 18 | 46.4 (41-53) | 11.1 | 27:73:00 | 10.3 (0-18) |
|  | IPD | 11 | 46.0 (39-68) | 45.4 | 64:36:00 | 17.0 (8-20) |
| Brain | sCJD | 51 | 59.15 (38-87) | 41.18 | 65:10:25 | 0 |
|  | Control | 33 | 74 (41-89) | 54.2 | Unknown | 0 |

sCJD (sporadic CJD); AD (Alzheimer's Disease); iCJD (iatrogenic CJD); IPD (inherited Prion Disease).

**Figure 1:**
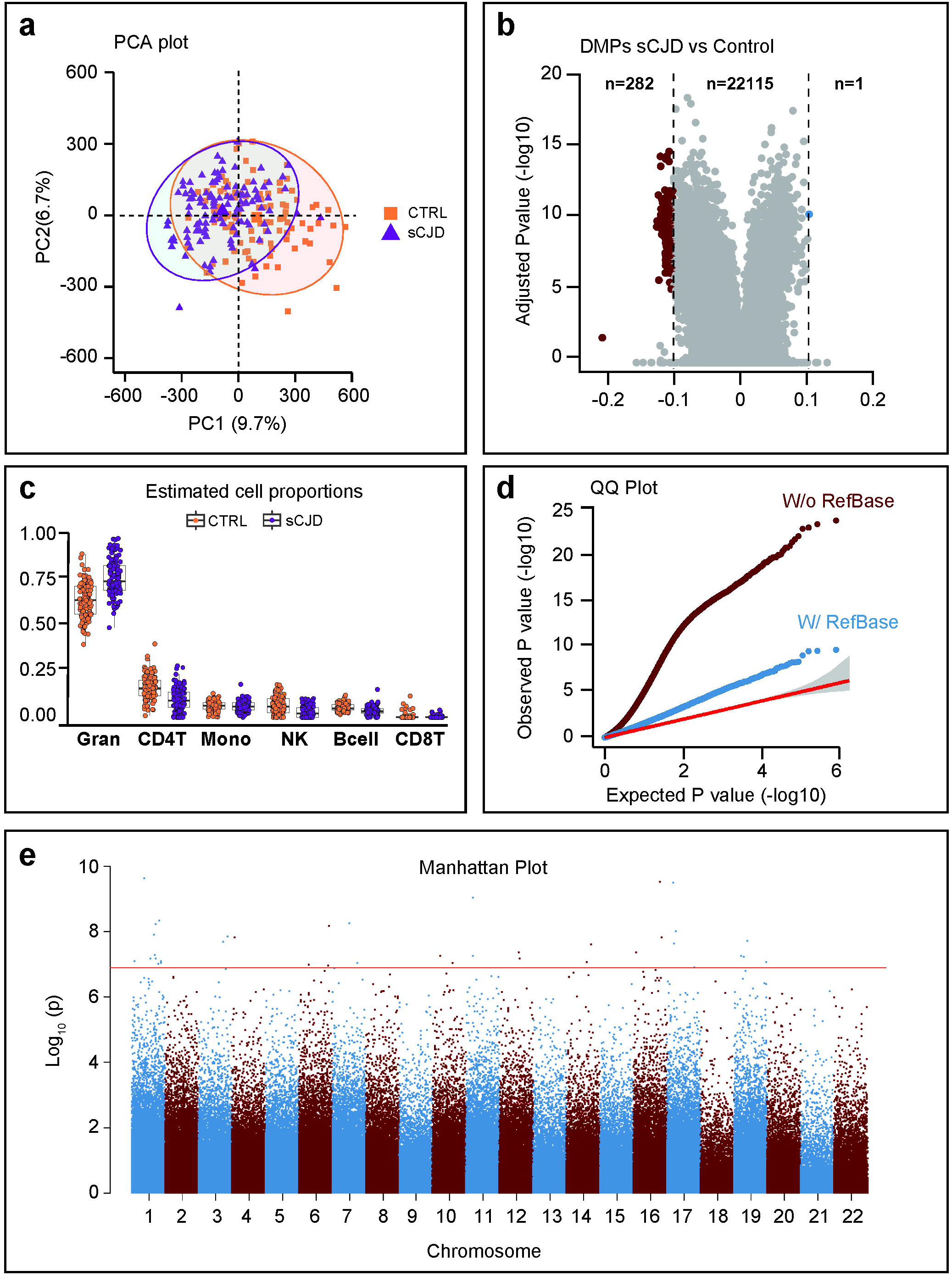
Genome-wide differential methylation in sporadic CJD blood. (**a**) Principal component analysis (PCA) of 219 DNA methylation profiles showing the first (PC1) and second (PC2) principal components (9.7% and 6.7% of the total variance). 95% confidence ellipses are drawn around the two groups: sCJD (triangles, purple) and healthy controls (squares, orange). (**b**) Volcano plot DMP association analysis (X and Y chromosomes were excluded from analysis), corrected for gender but not for blood cell composition. X-axis represents (effect size) adjusted mean delta difference, Y-axis represents-log10(q-value). Vertical lines indicate delta beta > |0,1|. (**c**) Tukey boxplots showing proportions of six different cell types as estimated by Houseman algorithm in sCJD (purple) and healthy controls (orange). Wilcoxon-Mann-Whitney test to identify differences between sCJD and control: Granulocytes p=1.47e-14; CD4T p=5.05e-11; Monocytes p=0.78; Natural Killers cells p=1.10e-08; B cell p=1.55e-10; CD8T p=0.59 (**d**) Quantile-quantile plots (QQ plots) of the distribution of observed-log10 association p values against the expected null distribution without (dark red) and with (blue) cell type correction. The red line represents the expected distribution with 95% confidence interval. Genomic inflation lambda scores are given in each QQ plot to quantify statistical inflation of p values. (**e**) Manhattan plot of probes associated with disease status corrected for blood cell type composition. Red line indicates significance threshold (Bonferroni-adjusted = 1.24×10^-7^). X-axis represents ranked chromosomes, Y-axis represents-log10 (p-value).

Given that whole blood is a heterogeneous collection of different cell types, each with their own distinct DNA methylation landscape, we next used Houseman’s statistical method to estimate the relative proportions of cell type components[22]. Figure 1 shows that cell proportion estimates (the sum of which is forced to 100%) differed subtly between samples in the study. The algorithm estimated that granulocytes comprised more than 75% of the blood cell types in 58 sCJD patients and 16 healthy controls, which is the upper limit of the normal range for granulocytes proportion in whole blood. When accounting for differences in the six cell types, the number of differentially methylated probes that passed genome-wide significance (p < 1.24×10-7) dropped from 22,398 to 38 (Supplementary Fig. 1c-e, online resource and Table 2). Strikingly, this cell type correction substantially reduced the inflation factor (λ) of our epigenome-wide analysis from 5.12 to 1.72 (Fig. 1d). Hierarchical clustering analysis (Pearson minus one correlation) of the significant 38 DMPs from the 219 patients and controls identified 3 clusters of sCJD cases and one cluster of controls (Supplementary Fig.1f) raising the possibility of heterogeneity in DNA methylation associated with the disease. In contrast to disease status, *PRNP* codon 129 genotype and sex of the patients did not cluster within the data. Of the significant 38 DMPs, 4 were hypomethylated with a mean effect size of Δβ-0.037 (95%CI±2.01×10^-2^), and 34 were hypermethylated with a mean effect size of Δβ +0.022 (95%CI±3.67×10^-3^). Figure 1 shows a Manhattan plot for these DMPs. Altogether, these results suggest sCJD patients have distinct blood DNA methylation profiles compared with healthy controls.

**Table 2.**
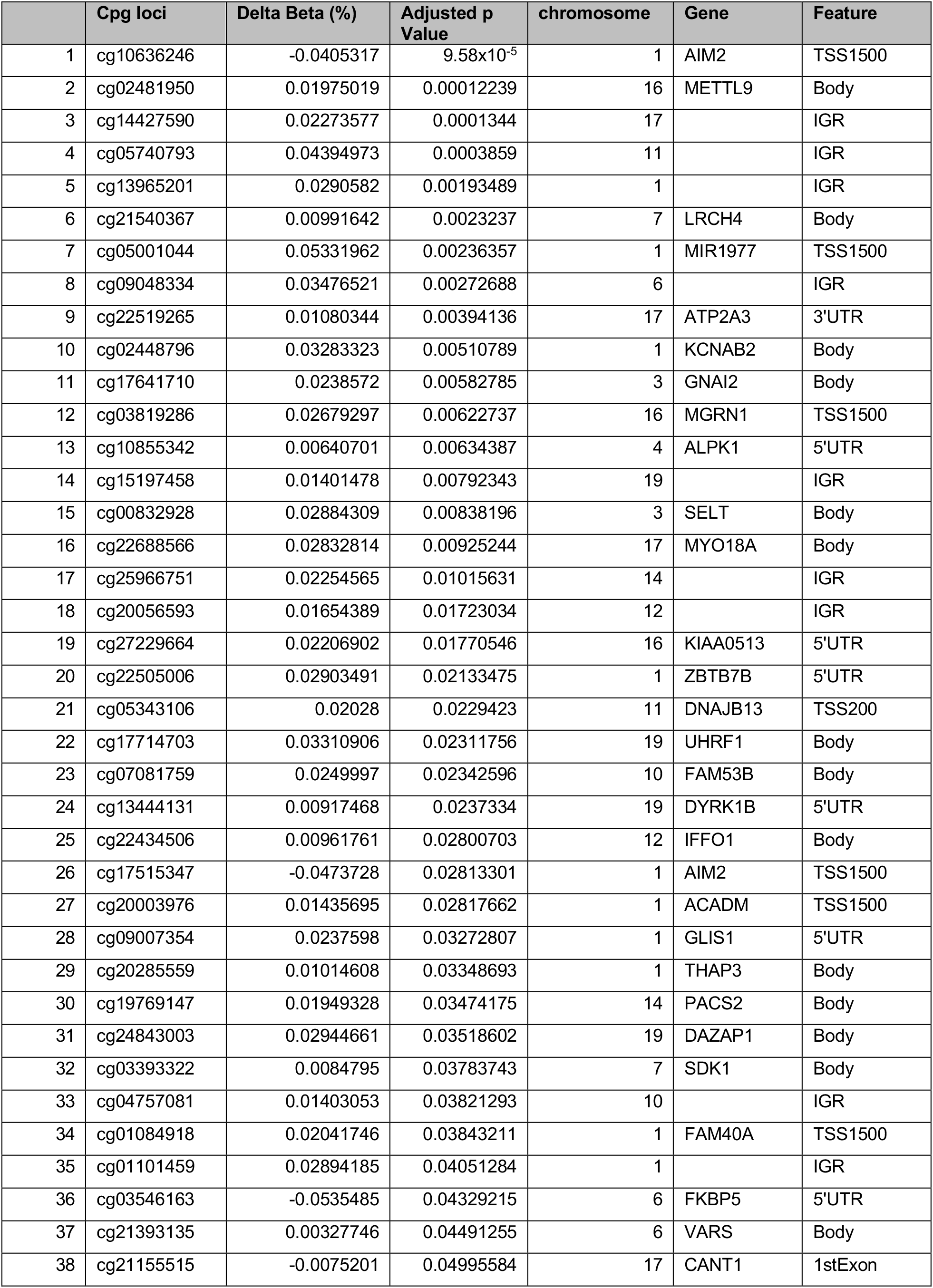
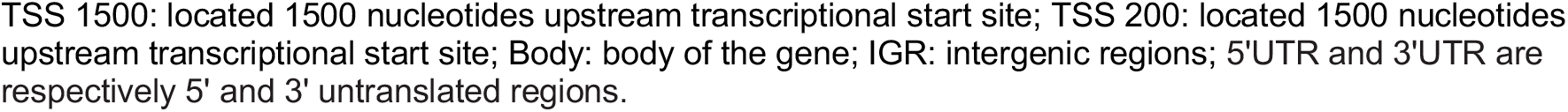
List of 38 Differentially methylated positions.

|  | Cpg loci | Delta Beta (%) | Adjusted p Value | chromosome | Gene | Feature |
| --- | --- | --- | --- | --- | --- | --- |
| 1 | cg10636246 | -0.0405317 | 9.58x10<sup>-5</sup> | 1 | AIM2 | TSS1500 |
| 2 | cg02481950 | 0.01975019 | 0.00012239 | 16 | METTL9 | Body |
| 3 | cg14427590 | 0.02273577 | 0.0001344 | 17 |  | IGR |
| 4 | cg05740793 | 0.04394973 | 0.0003859 | 11 |  | IGR |
| 5 | cg13965201 | 0.0290582 | 0.00193489 | 1 |  | IGR |
| 6 | cg21540367 | 0.00991642 | 0.0023237 | 7 | LRCH4 | Body |
| 7 | cg05001044 | 0.05331962 | 0.00236357 | 1 | MIR1977 | TSS1500 |
| 8 | cg09048334 | 0.03476521 | 0.00272688 | 6 |  | IGR |
| 9 | cg22519265 | 0.01080344 | 0.00394136 | 17 | ATP2A3 | 3'UTR |
| 10 | cg02448796 | 0.03283323 | 0.00510789 | 1 | KCNAB2 | Body |
| 11 | cg17641710 | 0.0238572 | 0.00582785 | 3 | GNAI2 | Body |
| 12 | cg03819286 | 0.02679297 | 0.00622737 | 16 | MGRN1 | TSS1500 |
| 13 | cg10855342 | 0.00640701 | 0.00634387 | 4 | ALPK1 | 5'UTR |
| 14 | cg15197458 | 0.01401478 | 0.00792343 | 19 |  | IGR |
| 15 | cg00832928 | 0.02884309 | 0.00838196 | 3 | SELT | Body |
| 16 | cg22688566 | 0.02832814 | 0.00925244 | 17 | MYO18A | Body |
| 17 | cg25966751 | 0.02254565 | 0.01015631 | 14 |  | IGR |
| 18 | cg20056593 | 0.01654389 | 0.01723034 | 12 |  | IGR |
| 19 | cg27229664 | 0.02206902 | 0.01770546 | 16 | KIAA0513 | 5'UTR |
| 20 | cg22505006 | 0.02903491 | 0.02133475 | 1 | ZBTB7B | 5'UTR |
| 21 | cg05343106 | 0.02028 | 0.0229423 | 11 | DNAJB13 | TSS200 |
| 22 | cg17714703 | 0.03310906 | 0.02311756 | 19 | UHRF1 | Body |
| 23 | cg07081759 | 0.0249997 | 0.02342596 | 10 | FAM53B | Body |
| 24 | cg13444131 | 0.00917468 | 0.0237334 | 19 | DYRK1B | 5'UTR |
| 25 | cg22434506 | 0.00961761 | 0.02800703 | 12 | IFFO1 | Body |
| 26 | cg17515347 | -0.0473728 | 0.02813301 | 1 | AIM2 | TSS1500 |
| 27 | cg20003976 | 0.01435695 | 0.02817662 | 1 | ACADM | TSS1500 |
| 28 | cg09007354 | 0.0237598 | 0.03272807 | 1 | GLIS1 | 5'UTR |
| 29 | cg20285559 | 0.01014608 | 0.03348693 | 1 | THAP3 | Body |
| 30 | cg19769147 | 0.01949328 | 0.03474175 | 14 | PACS2 | Body |
| 31 | cg24843003 | 0.02944661 | 0.03518602 | 19 | DAZAP1 | Body |
| 32 | cg03393322 | 0.0084795 | 0.03783743 | 7 | SDK1 | Body |
| 33 | cg04757081 | 0.01403053 | 0.03821293 | 10 |  | IGR |
| 34 | cg01084918 | 0.02041746 | 0.03843211 | 1 | FAM40A | TSS1500 |
| 35 | cg01101459 | 0.02894185 | 0.04051284 | 1 |  | IGR |
| 36 | cg03546163 | -0.0535485 | 0.04329215 | 6 | FKBP5 | 5'UTR |
| 37 | cg21393135 | 0.00327746 | 0.04491255 | 6 | VARS | Body |
| 38 | cg21155515 | -0.0075201 | 0.04995584 | 17 | CANT1 | 1stExon |

TSS 1500: located 1500 nucleotides upstream transcriptional start site; TSS 200: located 1500 nucleotides upstream transcriptional start site; Body: body of the gene; IGR: intergenic regions; 5'UTR and 3'UTR are respectively 5' and 3' untranslated regions.

### Characteristics of differentially methylated sites in sCJD

A circular ideogram was used to visualize the chromosome distribution of the 38 positions (DMPs) identified in sCJD patients’ blood (Fig. 2a and Supplementary Table 2, online resource, respectively). Genomic features of the 38 DMPs and 67 differentially methylated regions (DMRs) were compared to the null distribution of CpG probes included in the array and no significant difference was found (DMPs: Chi-square = 3.0267, df = 6, p-value = 0.8055; DMRs: Chi-squared= 204.64, df = 6, p-value < 2.2×10^-16^) (Supplementary Fig. 2a, online resource). Next, we asked if sCJD-specific DMPs were enriched for transcription factor binding motifs. Using PWMEnrich[54], we identified the most significantly over-represented motifs within the DMPs (Fig. 2b and Table 3) as those recognised by *GLTPD1*, a negative regulator of interleukin-1 beta secretion (p-value = 1.55×10^-5^). Other transcription factors identified included cell cycle regulators *DBP* and *DIABLO*, an inhibitor of apoptosis protein (IAP)-binding protein (p-value = 2.63 x10^-5^ and p-value = 0.0002, respectively). To further investigate potential consequences of the site-specific DNA methylation in sporadic CJD, we curated a list of RefSeq genes overlapping each of the 38 differentially methylated site and performed downstream analysis using Metacore Gene Ontology[37]. The results shown in Figure 2c show enrichment of negative regulators of protein tyrosine phosphatase activity, negative regulators of adenylate cyclase-activating adrenergic receptors, and negative regulators of cAMP-mediated signalling (Supplementary Fig. 2b, online resource).

**Figure 2:**
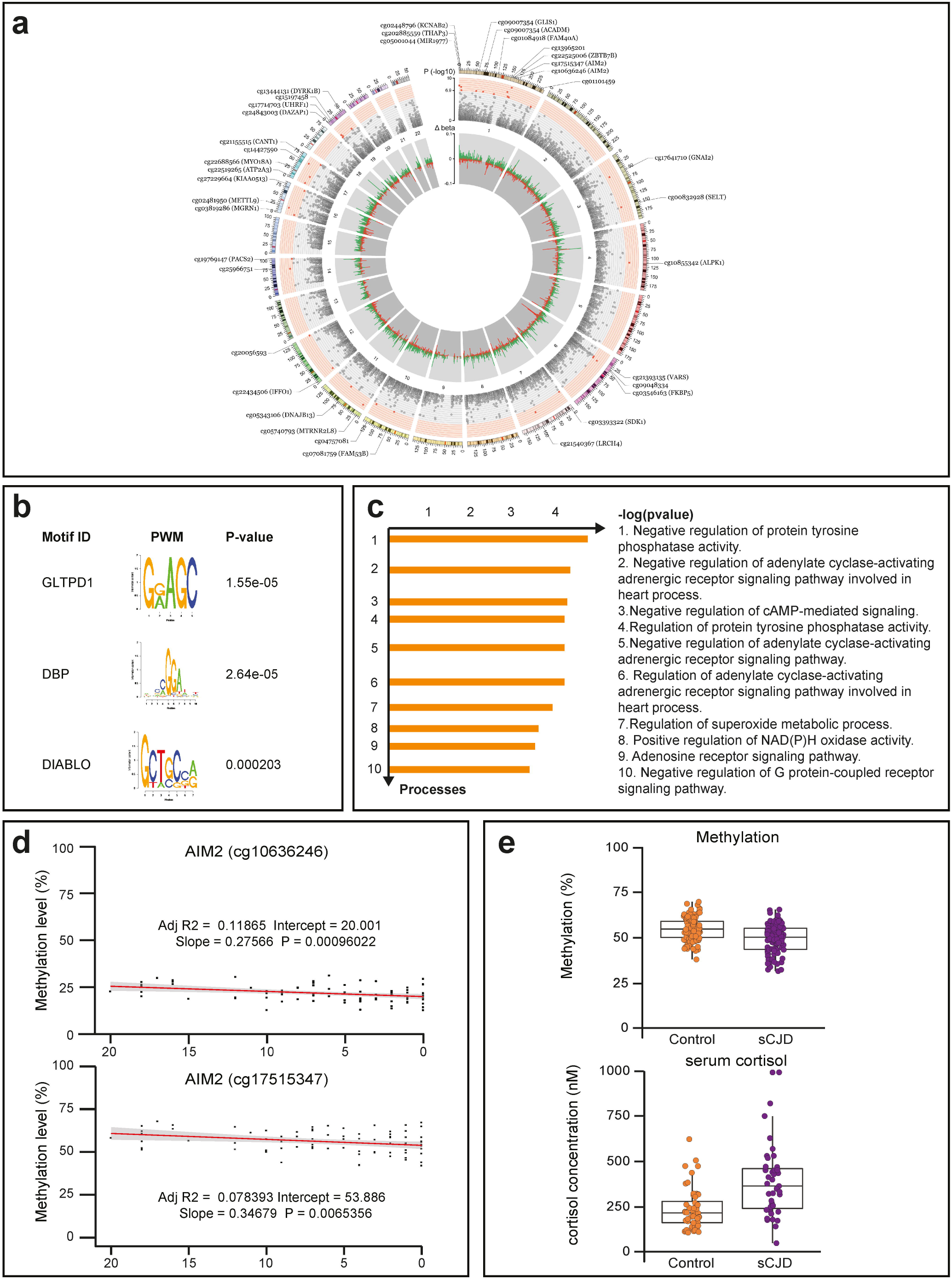
Key findings replicated using pyrosequencing in an independent case-control cohort. (**a**) Circos plot of epigenome-wide methylation levels in sCJD. Outermost circle represents the chromosome ideogram. Middle circle shows p-values (-log10) of the top 25,000 most significant DMPs (Bonferroni significance threshold of p<1.24×10-7 or –log10(p)>6.9 is distinguished with a red background). Significant DMPs are shown in red and labelled accordingly. The innermost circle represents the Δβ values across the genome, with hypermethylation in green and hypomethylation in red. (**b**) Top 10 motifs enriched in sequences within ±50 bp of significant DMPs within +/- 122 bases flanking the CG found under ‘Forward_Sequence’ heading in the Illumina HumanMethylation450 manifest file of the 38 significant DMPs. Ranking based on p-value. (**c**) Top 10 Gene Ontologies enriched in genes overlapping DMPs as identified using MetaCore (p-value threshold = 0.1). (**d**) Pearson coefficients between MRC Scale score with hypomethylation at two CpGs in the *AIM2* promoter (cg10636246 and cg17515347). (**e**) **Left**: DNA methylation levels (%) at *FKBP5* cg03546163 from the discovery study in sCJD patients (n=114; purple) and healthy controls (n= 105; orange) limma p=1.07e-07, corrected p=0.043. **Right**: serum cortisol concentrations (nM) in 39 sCJD patients (purple, 239.8 nM) and 52 controls (orange, 387.6 nM) (p = 6.6×10^- 5^).

**Table 3.** PMWEnrich motifs.

| Rank | Target | id | Raw Score | p.value |
| --- | --- | --- | --- | --- |
| 1 | GLTPD1 | MGC10334 | 1.927 | 1.55e-05 |
| 2 | DBP | M5338_1.02 | 1.37 | 2.63e-05 |
| 3 | UW.Motif.0555 | UW.Motif.0555 | 7.02 | 0.0001 |
| 4 | DIABLO | DIABLO | 2.46 | 0.0002 |
| 5 | EBF1 | M5364_1.02 | 1.43 | 0.0002 |
| 6 | ATF3 | M4683_1.02 | 5.54 | 0.0002 |
| 7 | UW.Motif.0654 | UW.Motif.0654 | 5.43 | 0.0002 |
| 8 | UW.Motif.0283 | UW.Motif.0283 | 15.02 | 0.0002 |
| 9 | CERS4 | LASS4 | 1.34 | 0.0003 |
| 10 | GOT1 | GOT1 | 1.14 | 0.0003 |

Top ten most position weight matrix (PWM) motifs found enriched in the 38 DMPs. The first column is the rank, the second shows the target name, the next column is the motif ID. This ID comes from the MotifDb package and can be used to look up further information about the motif. The fifth column gives the estimate p-value.

Using these 38 probes, we set out to examine whether patient metadata would help identify disease-modifying loci. We investigated the effect of *PRNP* codon 129 polymorphism [which is known to alter susceptibility to prion disease and rate of disease progression][35], age at onset and disease duration [binned in three groups: less than 100 days, longer than 200 days, or between 100 and 200 days] and found no significant difference between any of these groups (Supplementary Fig. 3a-c, online resource). Next, we explored disease severity using the MRC Scale, which rates functional impairment in sCJD from a score of 20 (healthy) to 0 (moribund)[58]. We correlated the patient’s score measured at the time each sample was collected with the methylation values at each DMP. The test was performed genome-wide (Supplementary Fig. 3d, online resource) and again with the 38 significant DMPs only (Fig. 2d). Figure 2 shows that methylation at two probes located in the promoter of *AIM2* (identified as a hit locus in the case-control study) decreased with disease progression (cg10636246: slope=0.27186, p-value = 0.0012915; cg17515347 slope=0.31729, p-value = 0.011766) whilst none of the other probes tested showed significant association (Supplementary Fig. 3e, online resource).

Given the previously reported link between *FKBP5* and neurodegeneration [5], our finding that the promoter of *FKBP5* is demethylated in sCJD (Fig. 2e and Table 2) prompted us to investigate the cortisol levels in sCJD patients. *FKBP5* acts as a cochaperone in modulating glucocorticoid receptor activity in the brain and periphery[62,64]. We analyzed cortisol levels in sera from 39 sCJD patients and 52 healthy controls. Figure 2 shows that cortisol levels were significantly elevated in sera from sCJD patients, with median cortisol concentration being 147.9 nM higher in sCJD patients (95% CI 77.8-218.1 nM, p-value = 6.6×10^-5^).

### The identified DNA methylation signature is unique to blood and sCJD

Next, we aimed at replicating these findings and explore the disease and tissue specificity of the associations. To this end we designed a second case/control study and determined the sample size required to power individual bisulphite pyrosequencing assays at each DMPs. An independent cohort of 72 sCJD and 114 age-matched controls was recruited (Table 1), and DNA methylation at candidate sites was profiled using pyrosequencing. Out of the 38 DMPs, 7 probes at 6 loci were selected for replication based on Bonferroni-adjusted statistical significance, effect size, and association with clinical metrics (MRC Scale score) observed in the discovery study (at or close to *AIM2* [cg10636246 and cg17515347], *FKBP5, METTL9, UHRF1, KCNAB2, MIR1977)*. Two more sites were used as controls: one within the prion gene *(PRNP)* and another within *ANK1*, a gene whose hypermethylation has consistently been reported in Alzheimer’s Disease (AD)[33]. DNA methylation levels at these 2 control sites remained unchanged in the discovery study. In total, 7 sites identified from the discovery study replicated (Fig. 3a): five DMPs and two controls sites. Given the length of the amplicon analyzed via the pyrosequencing assays, this allowed us to quantify another 5 CpG sites not present on the 450K array, adjacent to the tested DMPs in *FKBP5, AIM2*, and *UHRF1*, which also exhibited significant differential methylation between sCJD and healthy controls (Fig 3b). Altogether, the replication study identified a sporadic CJD methylation signature that comprises a total of 10 sites (5 DMPs from the array and 5 sites from the pyrosequencing) overlapping 4 genes *(AIM2, FKBP5, METTL9, UHRF1)*.

**Figure 3:**
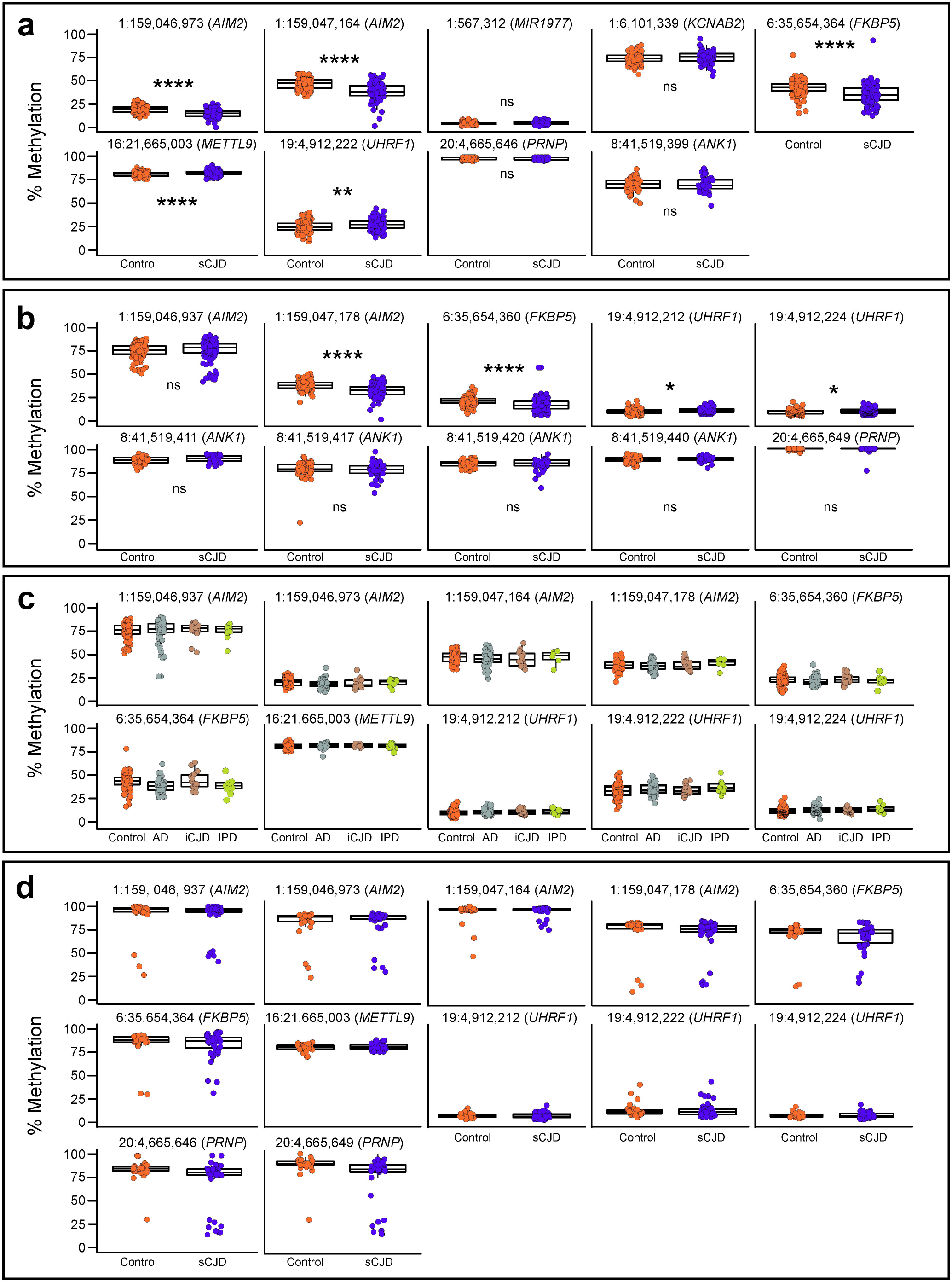
Differential methylation signature is unique to sCJD and to blood. (**a**) DNA methylation levels (%) at each DMPs chosen for replication by pyrosequencing sCJD patients (purple) and controls (orange). Labels above each plot show genomic coordinates and overlapping genes. (**b**) DNA methylation levels (%) at CpG sites adjacent to DMPs in sCJD patients (purple) and controls (orange). Labels above each plot show genomic coordinates and overlapping genes. (**c**) DNA methylation (%) at replicated DMPs in Alzheimer’s disease (grey), iatrogenic CJD (brown) and inherited prion disease patients (green) compared to controls (orange). (**d**) Methylation at replicated sites in frontal-cortex derived DNA from sCJD patients (purple) and non-demented controls (orange). See Supplementary Table 3, online resource, for all p-values. p-value<0.05 (*); p-value<0.01 (**); p-value<0.001 (***); p-value<0.0001 (****).

Next, we wanted to establish the disease-specificity of the DNA methylation signature in the context of the differential diagnosis of dementia. Blood samples from Alzheimer’s disease, iatrogenic CJD, and inherited prion disease patients were collected, and pyrosequencing was used to measure DNA methylation at the 10 sites (Supplementary Table 3, online resource). Figure 3 shows that none of the 10 sites showed significantly altered DNA methylation levels in any of the tested non-sCJD cohorts, suggesting the DNA methylation signature is specific to sCJD.

We investigated whether changes observed in sCJD blood reflected DNA methylation alterations in the brain. The same pyrosequencing assays were performed using frontal-cortex-derived DNA obtained from 51 sCJD patients and 33 non-cognitively impaired controls. Intriguingly, none of the sCJD-specific sites differentially methylated in blood were differentially methylated in brain (Fig. 3d). Altogether these results demonstrate that a DNA methylation signature identified in blood from sCJD patients replicates in an independent case-control cohort using a different technology and that the signal is not found in sCJD brain, or in the blood of other prion diseases or Alzheimer’s disease.

### DNA methylation array profiles to refine sCJD diagnosis and disease duration

Next, we sought to investigate whether DNA methylation changes could identify potential avenues for sCJD patient management by acting as diagnostic or prognostic biomarkers. To explore if DNA methylation array profiles could discriminate sCJD from healthy controls we applied a deep learning neural network classifier to our data. For each individual, the 1000 most significantly altered sites identified from the discovery study (114 sCJD and 105 controls) were selected and partitioned into training and test sets (50:50 ratio, with equal proportions of controls and patients in each set). Model accuracy varied during training, with an overall positive trend across sequential epochs. After 200 epochs validation accuracy appeared to plateau and the model started to overfit (Supplementary Fig. 3a-b, online resource). The model performed well in minimizing loss also called “binary cross-entropy loss”, indicating that the predicted probability converged to the actual label. After 10-fold cross-validation, the model had an accuracy of 87.24 (95%CI±3.16%) with an upper limit of 91.43% accuracy. As shown in Figure 4, the receiver operating characteristic (ROC) curve, the trained neural network model demonstrated a better performance compared to a basic Random Forest classifier with an AUC of 0.979 compared to 0.885, respectively (sensitivity 0.9074; specificity 0.8039). When trained using only the 38 significantly altered loci identified in the discovery study, the model achieved an accuracy of 79.45% (±1.09%).

**Figure 4:**
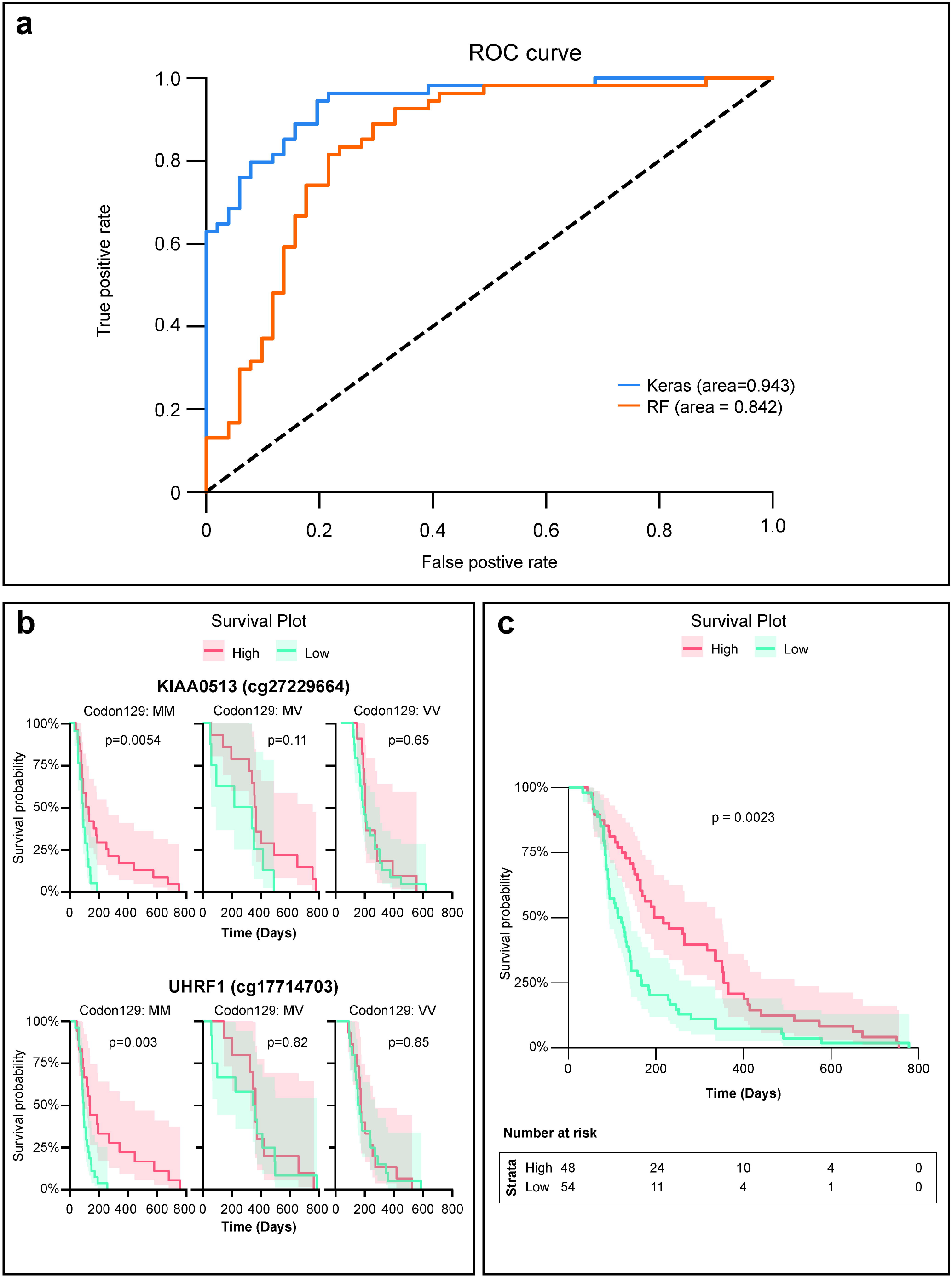
Diagnostic and prognostic utility of a DNA methylation in sCJD. (**a**) Receiver operating characteristic (ROC) curve performance comparison between neural network model (Keras; blue) and random forest classifier (RF; orange) on the validation set. (**b**) Kaplan-Meier survival analysis. Patients were divided into 3 groups based on the genotype at PRNP codon 129 (MM, VV or MV). Patients were divided into high (above median; red) and low (below media; green) DNA methylation values at *KIAA0513* and *UHRF1* DMPs. p-values were calculated using the log-rank test. (**c**) Survival analysis for 3 sites *(DNAJB13, GNAI2, UHRF1)* independent of PRNP genotype. Beta values from these 3 DMPs were transformed into z-scores. Kaplan-Meier curves using the average z-score (z-score >0, “High”; z-score <0, “Low”). See Supplementary Table 4, online resource, for median survival values.

Finally, we evaluated the association between DNA methylation levels and survival. In our cohort, as expected, codon 129 of the prion gene *PRNP* strongly impacts disease duration: MV heterozygous individuals had the longest disease duration whilst MM homozygous carriers die soonest (Supplementary Fig. 3c, online resource). To evaluate the influence of DNA methylation on survival, DNA methylation levels at each of the 38 DMPs were divided into high or low, based on the mean levels and correlated with disease duration. We found that elevated methylation levels at 8 DMPs (cg01084918 *[FAM40A]*, cg05343106 [DNAJB13], cg09007354 *[GLIS1]*, cg13965201, cg17641710 *[GNAI2]*, cg17714703 *[UHRF1]*, cg25966751 and cg27229664 *[KIAA0513])*, was associated with a longer survival in patients with sCJD (Supplementary Fig. 3d, online resource). When taking the genotype of *PRNP* codon 129 into account, this analysis revealed that the level of DNA methylation at two sites, *(UHRF1* and *KIAA0513)*, refines the prediction on disease duration for MM patients only (Fig. 4b). When not accounting for the effect of codon 129, 3 DMPs had an effect on survival: cg05343106 *(DNAJB13)*, cg17641710 *(GNAI2)*, and cg17714703 *(UHRF1)*. This effect was seen in the “Low” methylation group of cg05343106 *(DNAJB13)* and cg17641710 *(GNAI2)*, and “High” methylation group of cg17714703 *(UHRF1)*. Furthermore, combining together these 3 loci methylation profiles provided greater accuracy than each locus in predicting clinical outcomes. The distribution of beta values from these 3 DMPs were not significantly different from normality, thus we transformed them into Z-scores (Shapiro-Wilk cg01084918 p-value = 0.4967; cg05343106 p-value = 0.4372; cg17641710 p-value = 0.2549). Average z-scores were correlated (z-score >0, “High”; z-score <0, “Low”) with disease duration (Supplementary Table 4, online resource). Figure 4 shows that sCJD patients with higher levels of DNA methylation levels at those 3 sites had a 97 days longer median survival time, independently of *PRNP* genotype at codon 129. Together these results demonstrate a potential utility of profiling DNA methylation in whole blood from patients with sCJD: these profiles can help discriminate sCJD patients from sex and age matched healthy controls and may help predict disease duration.

## Discussion

Whilst DNA methylation has become increasingly studied in the context of neurodegenerative disorders, DNA methylation profiles have not yet been investigated in human prion diseases. Here, we performed a case-control study to analyse the relationship between DNA methylation and sporadic CJD using 405 peripheral blood samples from patients and controls. The discovery study used a genome-wide 450K Illumina BeadChip array, with replication using a second technology, pyrosequencing. Seven of nine sites that we identified successfully replicated. We went on to show that these effects were tissue and disease specific and could be exploited for diagnostic and biomarker purposes. Overall, we highlight the potential of DNA methylation array profiling of peripheral blood for a rare and serious neurodegenerative disorder.

Like GWAS, the more recently developed EWAS is subject to biases, in particular variability in the measured methylome differences between case and controls groups unrelated to the pathobiology of interest. Strategies have been developed to measure and correct for biases, in particular we found Houseman’s reference-based algorithm, that corrects for alterations in cell composition, proved successful in reducing the study-wide inflation[61] [22]. In this case, observed differences in the methylome might have resulted from a change in the cell types in blood that contributed to DNA in the study. The Housman algorithm estimates the relative proportion of major cell-types in blood samples, using validated methylation markers. No differences in sCJD blood cell types have previously been detected[31,8], although to date no large-scale studies have explored this aspect. The residual inflation of our discovery study (1.72) is higher than the lambda observed in similar studies[29] and raises the possibility that the false positive rate is not entirely controlled. In blood, comorbidity with systemic diseases and environmental factors such as nutrition [17] and environmental chemicals [21] have been linked to inflation in EWAS.

We questioned the relevance of the magnitude of the effect size we report here compared with the effects seen in cancerous tissues[50]. The mild to moderate effect size observed in sCJD blood could be caused by the fact that the signal is observed in the periphery rather than in the main tissue affected by the disease. To date, no genome-wide DNA methylation profiling has been reported in brain tissues in sporadic CJD. However, similarly mild effect sizes (around 10%) have been reported for significant DNA methylation changed observed in brain samples from other neurodegenerative disorders such as Alzheimer’s Disease (AD) [1], Multiple System Atrophy (MSA) [3], Amyotrophic Lateral Sclerosis (ALS) [18]. Another possible explanation for the mild effect size is that the difference in DNA methylation is cell-type specific, and therefore that the signal is diluted in whole blood. If this was the case, changes in DNA methylation at identified sites should be evaluated in different blood fractions. In line with this possibility is the fact that effect sizes are systematically greater in our replication study, where results were not corrected for cell type composition. Similarly, whether these DNA methylation alterations become magnified over the course of disease remains to be investigated as we did not test patients longitudinally. We replicated our findings in an independent case-control cohort of blood samples using an independent technology, namely pyrosequencing. Furthermore, differentially methylated sites in sporadic CJD patients remained unaffected in blood from other neurodegenerative disorders (iatrogenic CJD, inherited prion diseases, Alzheimer’s disease). One possible explanation for this is the fact that sCJD patients are generally at an advanced neurological state at diagnosis and when blood samples taken compared to other prion diseases and AD. To date, it remains difficult to confidently identify loci that replicate across studies given the few, relatively small EWAS studies in neurodegenerative diseases, their design and number of samples analysed [16].

Concordance of DNA methylation signatures between blood and brain has been reported in ALS[18]. However, the vast majority of the literature suggests that the degree of cross-tissue correlation for DNA methylation signals is not very high[6]. Studies in AD, Parkinson’s Disease (PD), and Huntington Disease (HD) all show very little overlap between blood and brain DNA methylation signals [6,14,20]. Our findings that DNA methylation profiles in peripheral blood do not mirror those in frontal cortex in sCJD corroborate these reports. Of note, these samples did not belong to the same individuals. Whether this general lack of correlation is due to the nature of the samples that are being compared (bloods are taken from living individuals whilst brain samples are collected post-mortem) requires further investigation.

Although blood-based DNA methylation might not be generally considered as a surrogate for brain tissues, DNA methylation profiles detected in peripheral tissues might remain useful as biomarkers[16]. Our findings support this. First, we report that loss of DNA methylation at 2 sites on the *AIM2* promoter correlates with disease severity. AIM2 is a key component of the inflammasome pathway, a component of the innate immune system that drives the production of the inflammatory cytokine interleukin-1β (IL-1β) in response to microbial and nonmicrobial signals [52]. In yeast, AIM2 triggering induces a prion-like polymerization of ASC into filaments that provide platforms for activating inflammatory cytokine production [7] [32]. However, prion pathogenesis does not seem to lead to inflammasome activation in mice[44]. Second, we show that sCJD patients display a concomitant decrease in *FKBP5* DNA methylation and elevated cortisol levels. FKBP5 binds to glucocorticoid receptors and modulates glucocorticoid sensitivity. Epigenetic regulation of FKBP5 and its consequences on patient’s behaviour is well documented: accelerated age-related decreases in FKBP5 methylation are associated with childhood trauma and depressive phenotypes [63] whilst increased DNA methylation levels of FKBP5 have been found in patients suffering from post-traumatic stress disorders (PTSD) and major depressive syndromes[27]. Such changes are reminiscent of the alterations observed in sCJD. Although additional functional work is needed to clarify the relationship between sCJD, FKBP5 and the hypothalamic pituitary adrenal (HPA) axis [where FKBP5 plays a major role], it could be that the HPA axis provides a link between the pathology in the brain and the periphery. Several lines of evidence support the implication of FKBP5 in prion diseases. Work from the Soto group showed that FK506, a Calcineurin (CaN) inhibitor which binds to FKBP5, substantially decreased the severity of clinical signs in mice presenting symptoms of prion disease. In the same study, the authors report that FK506, also known as Tacrolimus, reduces brain degeneration and increases survival[40]. Another study by Nakagaki et al demonstrated that FK506 markedly reduced the abnormal form of prion protein in the cell cultures[42]. Stocki et al proposed that FK506 treatment results in a profound reduction in PrP^C^ expression due to a defect in the translocation of PrP^C^ into the endoplasmic reticulum with subsequent degradation by the proteasome[53]. More recently, treatment with FK506 suppressed typical sCJD pathology (gliosis) and significantly prolonged the survival of sCJD-inoculated mice[41]. Finally, *FKBP5* DNA methylation decreases along the lifespan; this age-related decrease is not confounded by blood cell type heterogeneity and occurs in purified immune cell subtypes[63]. The same group also showed that FKBP5 upregulation promotes NFkB related peripheral inflammation and chemotaxis. The role played by FKBP5 in inflammation and PTSD resonates with the function of another gene found to be differentially methylated in sCJD blood: AIM2. Indeed, PTSD cases are more likely to have high levels of C-reactive protein (CRP), a widely-used measure of peripheral inflammation, and this association is mediated by methylation at the AIM2 locus[38].

We acknowledge several limitations of this study. Having independent replication and a larger sample size of DNA methylation data would further increase the generalizability of the classifiers identified in this study. Additionally, since methylation values can change throughout lifespan, it will be insightful to evaluate the methylation signature over the course of the disease, longitudinally. In the long-term, we seek to determine when the alterations to DNA methylation patterns begin to show relative to the onset of prion diseases, and whether robust and stable DNA methylation patterns can predict the onset of disease in those at high risk of the disorder. We also aim at investigating DNA methylation profiles in first degree relatives of the sCJD patients. We were underpowered to confidently determine whether changes discovered in sCJD were shared with other very rare prion diseases, so it remains possible our findings associate with multiple aetiological groups. Further, whilst we compared to non-prion disease neurodegenerative disorders, these affect more specific brain functions in early stages, and are less aggressive in progression. These superficial disease-related differences may contribute to observed differences in blood DNA methylation, not necessarily only pathways specific to prion disease pathobiology. Finally, the brain samples were not selected because of PrP abnormalities, yet all cases had immunohistochemistry performed qualitative reports have been produced.

What are the functional consequences of these mild changes for cellular malfunction and disease? Future work will help investigate the mechanisms that underlie role of the selected CpG on sCJD establishment and progression. In line with this, our study can only provide correlative evidence for a blood-based sCJD-specific DNA methylation signature that robustly discriminates sCJD patients from controls, and other types of prion disease and AD patients. EWAS studies do not allow us to infer whether the DNA methylation observed represents a cause or consequence of sporadic CJD. Further investigations and functional studies will be required to understand better the contribution of epigenetic changes to sCJD.

Finally, our results suggest that DNA methylation profiling could be of use to refine sCJD disease management. We show that a data-driven machine learning approach using DNA methylation profiles accurately distinguishes sCJD patients from healthy individuals using peripheral blood. To date, our study is the first to report an assay that is capable of identifying sCJD patients from a blood sample, and that discriminates between sCJD and AD. Moreover, it is also the first study to suggest that DNA methylation could be used as a blood biomarker in human prion diseases. In line with this, we additionally demonstrated that a risk score based on DNA methylation of three identified sites predicts disease duration. Work from Zhang et al. recently demonstrated that DNA methylation age acceleration associated with ALS survival [65]. The finding that DNA methylation levels on two sites *(KIAA0513* and *UHRF1)* refined survival information driven by *PRNP* codon 129 genotype is informative, particularly for individuals most at risk. PRNP codon 129 Methionine homozygosity is associated with shorter disease duration in sCJD [36] and therefore combining genetic and epigenetic information provides further insights than genotype only. None of these sites overlapped with established aging-related CpGs. Future work in independent cohorts of sCJD patients is needed before these methods might be considered for clinical use.

Regardless of the underlying mechanisms, our results demonstrate that non-protein mediated information about sCJD disease status is present in blood and suggest that mapping such DNA methylation patterns alterations, with future independent replication, might be of use for testing and counselling. The fact that there is a signal in sCJD blood is it not surprising when sCJD is thought to arise from spontaneous prion production in the brain as a stochastic event. Future work will unravel whether DNA methylation is also altered in acquired prion diseases, which involve peripheral pathogenesis and spread. This study is meaningful in providing new avenues for understanding sporadic CJD disease mechanisms and identifying biomarkers which complement existing clinical signals in the periphery.

## Data Availability

Data is available upon request

## Acknowledgements

We are extremely grateful to Prof Sebastian Brandner and Dr Zane Jaunmuktane for their continued support and advices. We thank all our colleagues and patients who have participated in the National Prion Monitoring Cohort Study and provided blood samples since 2008. We would like to acknowledge the NHS National Prion Clinic Team and Joanna Field for assistance with collation of patient metadata; Gaganjit Madhan Kaur and Mark Kristiansen from UCL Genomics for performing the array hybridisation. We wish to also thank Ankur Chakravarty, Rebecca Smith, and Prof Stephan Beck for helpful advice and technical assistance. We are very grateful to Richard Newton for his help with figures and graphics. The study was funded by the Medical Research Council (UK), Alzheimer’s Research UK, the NIHR Queen Square Dementia Biomedical Research Unit and the NIHR Biomedical Research Centre at University College Hospitals NHS Foundation Trust. We are very grateful to patients and volunteers who have donated blood samples and their time to research and have made this study possible. LD is supported by a Leonard Wolfson Foundation PhD fellowship. FG is supported by a NIHR Department of Health grant.

## Author Contributions

LD, SM and EV designed experiments. LD, FG and TC performed experiments. LD, FG, SM and EV interpreted data. SM and JC assessed patients and clinical parameters. LD and FG conducted bioinformatic and statistical analyses. TB, PS, AS, RS and KL provided technical support; RR and JMS provided samples. LD, SM, and EV wrote the manuscript. All authors approved the manuscript.

## Competing Interests statement

Prof Collinge is a director and shareholder of D-Gen Limited (London), an academic spinout company working in the field of prion disease diagnosis, decontamination, and therapeutics. None of the other authors report any conflict of interest.

## Notes

### Author Declarations

Ethical approval was obtained from the National Hospital Local Research Ethics Committee.

